# Insulin-like Growth Factor-Binding Protein 2 and Adverse Left Ventricular Remodeling After First Myocardial Infarction

**DOI:** 10.64898/2026.03.04.26347626

**Authors:** Meyer Elbaz, Marie-Hélène Grazide, Vincent Bataille, Grégoire Blanc, Paul Gautier, Rodwell Mkhwananzi, Hueseyin Firat, Cécile Vindis

## Abstract

**Background and Aims:** Despite advances in reperfusion and medical therapy, survivors of acute myocardial infarction (AMI) remain at risk for adverse left ventricular remodeling (LVR), a precursor to heart failure. Building on prior work outlining 12-month biomarker trajectories linked to early ventricular dysfunction, we aimed to assess whether these circulating biomarkers predict long-term adverse LVR.

**Methods:** We prospectively enrolled 155 patients experiencing their first AMI. Clinical, biochemical, and echocardiographic data were obtained at pre-percutaneous coronary intervention (pre-PCI), 24 h post-PCI, discharge (day 3), 6 months, and 12 months. Adverse LVR was defined as an increase of ≥15 % in left ventricular end-systolic volume at 12 months.

**Results:** Adverse LVR occurred in 34 % of patients and was associated with cardiometabolic dysregulation (higher glucose, triglycerides, BMI, HOMA-IR; lower HDL-C). Among the six baseline biomarkers, only insulin-like growth factor-binding protein 2 (IGFBP-2) differed significantly between groups (p = 0.021) and remained independently associated in multivariable analysis (p = 0.036). Inclusion of IGFBP-2 increased the predictive model’s area under the receiver-operating characteristic curve from 0.735 to 0.801.

**Conclusions:** IGFBP-2 is an independent predictor of adverse LVR following AMI, highlighting the interplay between metabolic dysfunction and maladaptive remodeling. Incorporating IGFBP-2 into clinical risk models could improve stratification and guide precision therapies for high-risk patients.

## Introduction

Acute myocardial infarction (AMI), a major manifestation of coronary artery disease (CAD), remains a leading global health concern, contributing substantially to long-term mortality and morbidity. Despite advances in reperfusion strategies—particularly primary percutaneous coronary intervention (PCI)—and guideline-directed pharmacotherapy, AMI survivors remain at risk for recurrent infarction, heart failure (HF), and sudden cardiac death, with 10-year mortality rates approaching 30% (1).

Development of HF after AMI is closely linked to maladaptive left ventricular remodeling (LVR), which occurs in 5–10% of patients within the first year, even with timely PCI and optimal therapy (2). Over the past decade, pharmacological management of HF post-AMI has expanded beyond traditional cornerstones—β-blockers, angiotensin-converting enzyme (ACE) inhibitors, and mineralocorticoid receptor antagonists (MRAs) as foundational therapies (3)—to include sodium– glucose cotransporter 2 inhibitors (SGLT2i) and glucagon-like peptide-1 receptor agonists (GLP-1 RA), both of which confer cardiovascular benefit irrespective of diabetes status (4-6). This widening therapeutic landscape underscores the need for refined risk stratification tools to guide individualized treatment strategies.

Biomarkers of inflammation, fibrosis, and metabolism are increasingly recognized as potential companions in predicting adverse remodeling and informing therapeutic decisions. In our recent prospective study, we profiled the 12-month trajectories of a panel of circulating biomarkers in first-AMI patients (7). The selected biomarkers were chosen for their established association with left ventricular (LV) dysfunction, specifically reflecting key pathological processes such as fibrosis, remodeling, inflammation, and cellular infiltration at various disease stages. Among these biomarkers, baseline levels of soluble suppression of tumorigenicity 2 (sST2), interleukin-6 (IL-6), osteopontin, angiopoietin-2, growth differentiation factor 15 (GDF-15), and insulin-like growth factor-binding protein 2 (IGFBP-2) have been shown to correlate with N-terminal pro–B-type natriuretic peptide (NT-proBNP) concentrations and left ventricular ejection fraction (LVEF), thereby emerging as potential early indicators of post–AMI dysfunction. Building on these observations, the present study aimed to further examine the relationship between these circulating biomarkers and long-term adverse LV remodeling (LVR) in patients experiencing a first-time AMI.

## Methods

### Study Population

This prospective, longitudinal cohort study was conducted at the Toulouse University Hospital Center in France, involving patients admitted to the intensive care unit for AMI between September 2018 and July 2021 as previously described. The study protocol adhered to the ethical guidelines of the 1975 Declaration of Helsinki and Toulouse Hospital guidelines. It was reviewed and approved by the National Institution’s ethics committee on research on humans (Comité de Protection des Personnes, France), with approval number 2017-A01959-44. All patients included have signed an informed consent to participate in study.

Inclusion criteria were patients ≥18 years old hospitalized for a first AMI in the context of acute coronary syndrome, as defined by the Third Universal Definition of MI (8), and treated with PCI. Exclusion criteria comprised: resuscitation after cardiac arrest by methods other than single defibrillation; presence of cardiogenic shock; severe non-cardiac comorbidities with an expected survival <1 year; planned major non-cardiac surgery; pre-existing left ventricular dysfunction; any history of left-, right-, or biventricular heart failure; clinical evidence of significant heart failure on admission (Killip class ≥2); presentation >24 hours after symptom onset; or prior MI. The primary clinical outcome was the proportion of patients with adverse LVR as defined as a ≥15% change in LV end-systolic volume (LVESV) during 12-month follow-up (9, 10). This threshold aligns with established echocardiographic criteria, which commonly define post-MI remodeling as a ≥20% increase in left ventricular end-diastolic volume (LVEDV) or ≥15% increase in LVESV. Compared with LVEDV or ejection fraction changes, LVESV provides a more stable and reproducible measure of remodeling, particularly in longitudinal assessments (9, 10). Among the 119 patients who completed the study, 34 % (n=41) presented adverse LVR as defined. Based on the presence of remodeling at follow-up, the study population was divided into adverse LVR and no-adverse LVR patients.

### Measurement of biomarkers and clinical data collection

Baseline clinical data, laboratory measurements, and echocardiographic assessments were retrieved from electronic medical records at predefined time points: before PCI (H0), 24 hours after PCI (H24), at hospital discharge (D3), and during follow-up at 6 months (M6) and 12 months (M12). Transthoracic echocardiography (TTE) was specifically conducted at D3, M6, and M12. For each patient, plasma samples collected at the specified time points were centrifuged, aliquoted, and stored at ™80°C. The concentrations of sST2 were determined using the Aspect-Plus ST2 lateral flow immunoassay (Critical Diagnostics, San Diego, USA), following the manufacturer’s specifications. IGFBP-2, angiopoietin-2, osteopontin and IL-6 were measured using the Ella Automated Immunoassay System (Bio-Techne Corporation, Minneapolis, USA), adhering to the manufacturer’s assay characteristics. Routine biomarkers were assessed in the Biochemistry Laboratory of the Toulouse University Hospital Center. High-sensitivity cardiac troponin T (hs-cTnT), N-terminal pro-B-type natriuretic peptide (NT-proBNP), glucose and GDF-15 levels were measured in plasma aliquots using specific Elecsys® quantitative sandwich electro-chemiluminescence immunoassays on a Cobas 8000 analyzer (Roche Diagnostics, Mannheim, Germany), according to the manufacturer’s guidelines. Plasma insulin levels were determined using the insulin ELISA kit (Mercodia AB, Uppsala, Sweden) following the manufacturer’s specified assay characteristics. The homeostasis model assessment index of insulin resistance (HOMA-IR) was defined as fasting insulin x fasting glucose/22.5 (11). Creatinine was measured using the CREP2 (creatinine plus ver.2 test), and C-reactive protein (CRP) levels were determined using the Tina-quant CRP IV test, both conducted on a Cobas 8000 analyzer (Roche Diagnostics, Mannheim, Germany), following the manufacturer’s specified assay characteristics. Transthoracic echocardiography was performed in all patients using commercially available ultrasound systems (GE Vivid E9) in accordance with the recommendations of the American Society of Echocardiography and the European Association of Cardiovascular Imaging (12). Examinations were conducted within 48–72 hours of hospital admission for MI, and subsequently at follow-up time points (6 and 12 months) as per study protocol. Pre-PCI transthoracic echocardiograms are of limited diagnostic value, as these rapid, exploratory examinations primarily serve triage purposes; structural myocardial alterations—such as infarct expansion and LVR— typically occurs in the days to weeks after MI, not during the initial periprocedural window (13). Standard parasternal long-axis, short-axis, and apical views were obtained to assess LV size, wall motion, and systolic function. LVEF was calculated using the modified Simpson’s biplane method. Additional parameters included left ventricular end-diastolic and end-systolic volumes (LVEDV, LVESV), left atrial volume index (LAVI), transmitral Doppler inflow velocities (E and A waves), and tissue Doppler imaging (TDI) of mitral annular velocities (e⍰) to estimate diastolic function. Right ventricular function was assessed using tricuspid annular plane systolic excursion (TAPSE) and right ventricular fractional area change (RV FAC). Speckle-tracking echocardiography was performed offline to assess global longitudinal strain (GLS) using vendor-specific software. GLS was measured from apical views (four-, two-, and three-chamber), and a value of > –18% was considered normal based on published reference standards. To assess reproducibility, a random subset of 20% of echocardiographic studies was independently analysed by a second experienced cardiologist blinded to clinical data and primary readings. Interobserver and intraobserver variability were assessed by three independent observers using intraclass correlation coefficients for key parameters, including LVEF and GLS, with variability of approximately 5–8% for LVEF and 5–7% for GLS, in line with values typically reported in echocardiographic studies.

### Statistical analysis

Continuous variables are expressed as mean ± standard deviation (SD) or median (interquartile range [IQR]) depending on their distribution. Categorical variables are presented as counts and percentages. Comparisons between groups (adverse vs. non-adverse LVR) were performed using the Student’s t-test or Mann–Whitney U test for continuous variables and chi-squared or Fisher’s exact tests for categorical variables, as appropriate. A multivariable logistic regression model was used to identify factors independently associated with adverse LVR. Variables included in the initial model were selected based on clinical relevance and univariate significance (p < 0.15). The final model was obtained using a backward stepwise selection procedure, with variables retained based on a significance threshold of p < 0.05. To assess the incremental value of additional biomarkers, we compared nested logistic regression models using likelihood ratio (LR) χ^2^ tests. Alongside, we compared the discriminative performance of the models using ROC curves. Statistical significance was set at a two-tailed p < 0.05. Analyses were performed using Stata Statistical Software (StataCorp. 2023. Stata Statistical Software: Release 18. College Station, TX: StataCorp LLC).

## Results

### Population characteristics

We assessed a cohort of 155 patients who experienced a first AMI between September 2018 and July 2021, all of whom underwent PCI. The cohort had a mean age of 61.4 ± 10.8 years, with 80.7% male and 69% presenting with ST-elevation myocardial infarction (STEMI). The median LVEF on admission was 55% [IQR 51–60] (7). Of these, 119 patients completed the study and were included in the final analysis. Exclusion from the analysis was due to missed intermediate visits primarily related to disruptions caused by the COVID-19 pandemic. The baseline characteristics of the study population according to LVR status (adverse vs non-adverse) are presented in Table 1. Among the study participants, 34 % exhibited LVR at 12-month follow-up. The mean change in LVEF at 12-month follow-up (ΔLVEF) was significantly superior in the non-adverse LVR group (+6.2) compared to the adverse LVR group (+0.2; p < 0.001). The adverse LVR group presented significantly higher levels of blood glucose and triglycerides, as well as lower levels of high-density lipoprotein-cholesterol (HDL-C) and body mass index (BMI) (>27). The median Homeostasis Model Assessment of insulin resistance (HOMA-IR) index was higher in the adverse LVR group (4.1; interquartile range: 2.0–8.4) than in the no-adverse LVR group (2.7; interquartile range: 1.9–4.6), with a trend toward statistical significance (p = 0.062). These data show that patients who developed adverse LVR following AMI were more likely to exhibit a metabolic profile characteristic of insulin resistance and dyslipidemia. The concentrations of circulating sST2, IL-6, osteopontin, angiopoietin-2, GDF-15, and IGFBP-2, measured at baseline and at 12 months, were compared between the two study groups. As shown in Table 1, only IGFBP-2 levels differed significantly between patients with and without adverse LVR (p _baseline_ = 0.021; p _12-months_ = 0.035).

**Table 1:**
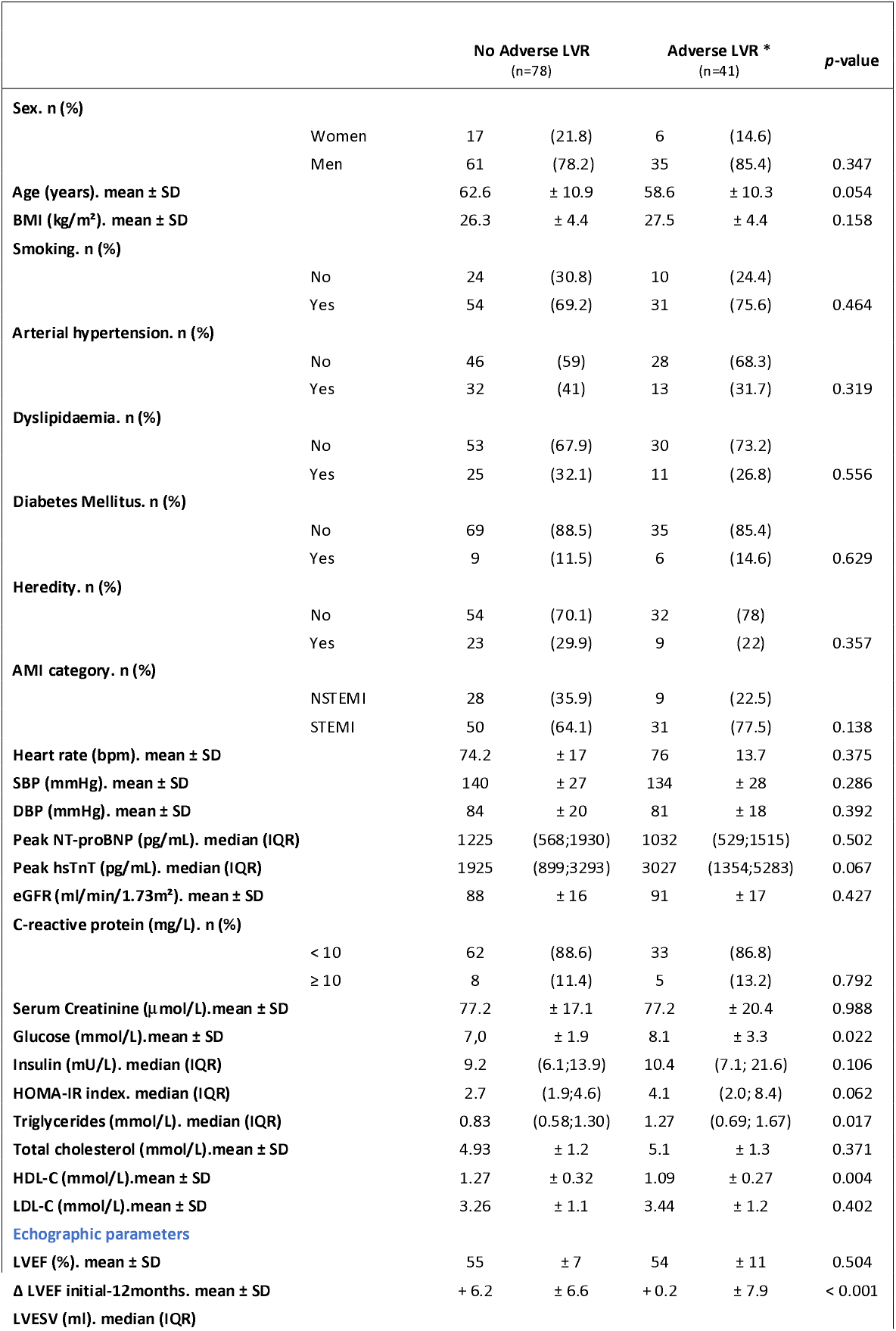
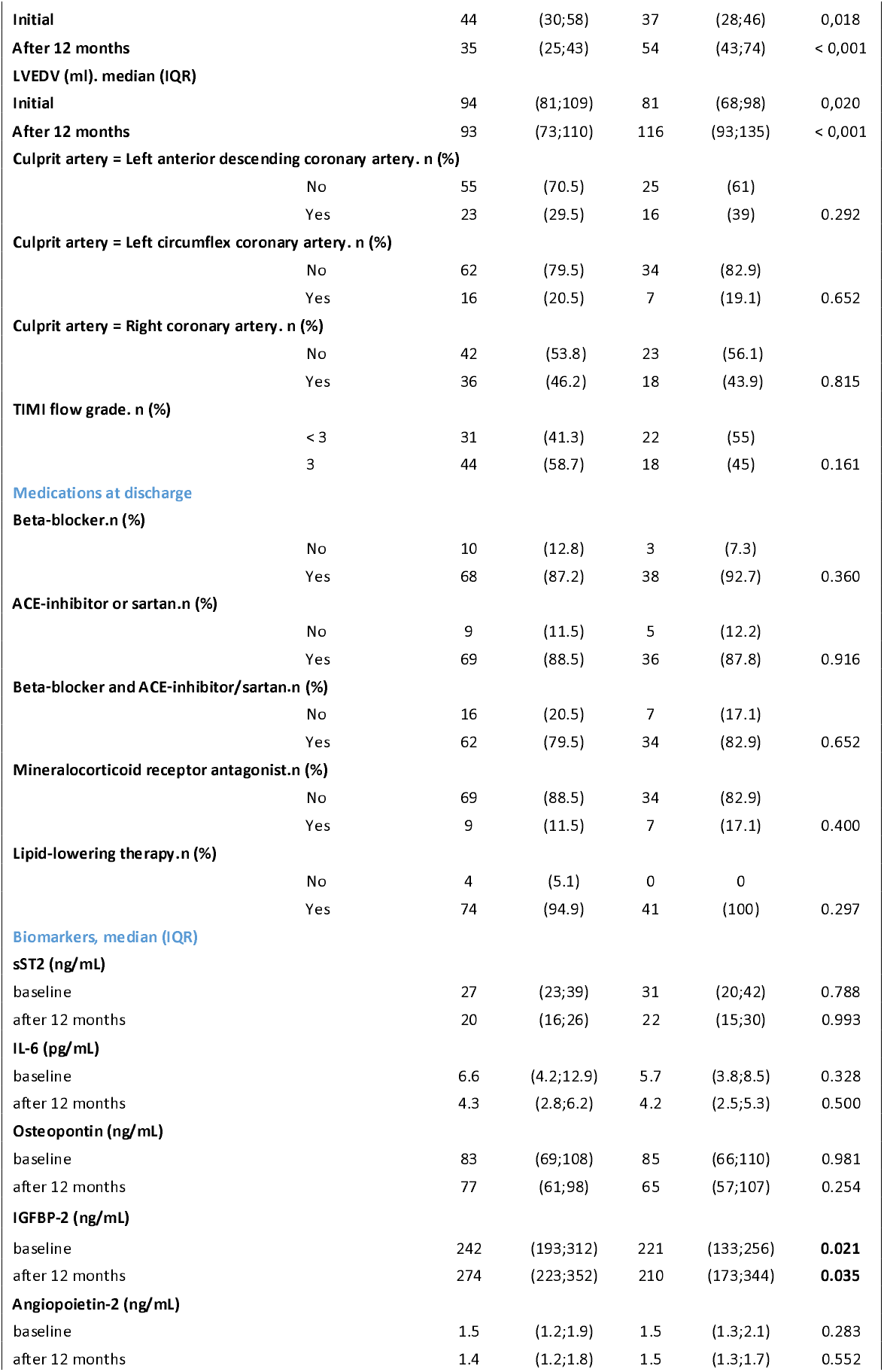
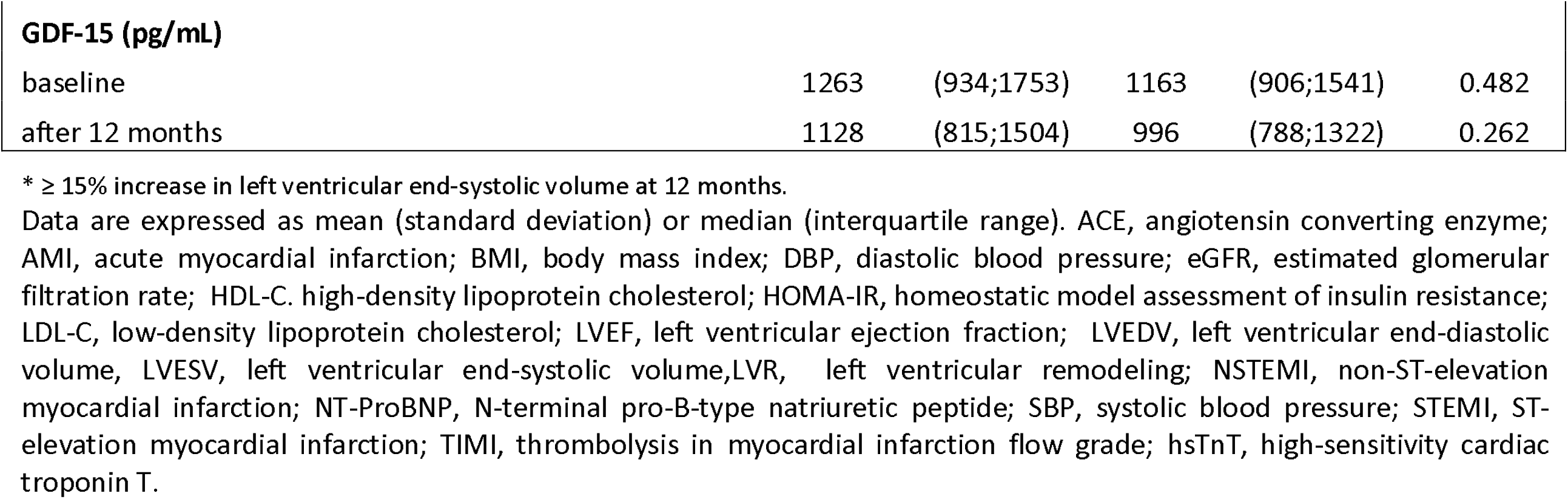
Baseline characteristics according to LVR status.

Proportion of patients exhibiting adverse LVR across stratified IGFBP-2 levels and association of IGFBP-2 with adverse LVR Patients who developed adverse LVR had significantly lower baseline IGFBP-2 levels (221 ng/mL) compared with the no-adverse LVR group (242 ng/mL; p = 0.021), as shown in Figure 1A. The incidence of adverse LVR was further examined across tertiles of IGFBP-2 concentrations (Figure 1B). The highest percentage of patients with an adverse LVR was found in the lowest tertile of IGFBP2 (<155 ng/mL), suggesting a potential association between reduced IGFBP-2 levels and unfavorable post-MI remodeling outcomes. Table 2 presents the results of the multivariable logistic regression analysis assessing the association between baseline IGFBP-2 levels and the risk of adverse LVR. After adjustment for ΔLVEF (the change in LVEF between baseline and 12 months) and HDL-C, IGFBP-2 levels remained independently associated with adverse LVR (p = 0.036), supporting the prognostic value of baseline IGFBP-2 in the post-MI setting.

**Table 2:** Multivariable logistic regression analysis for the risk of adverse LVR.

|  | **Odds-Ratio** | **95% CI** | ***p*-value** |
| --- | --- | --- | --- |
| **12 months LVEF evolution (%)** | 0,88 | 0,81 - 0,94 | < 0,001 |
| **HDL-C (mmol/L)** | 0,09 | 0,01 - 0,63 | 0,036 |
| **IGFBP-2 (ng/mL) \*** | 0,31 | 0,10 - 0,93 | 0,036 |

\* log-transformed.

**Figure 1.**
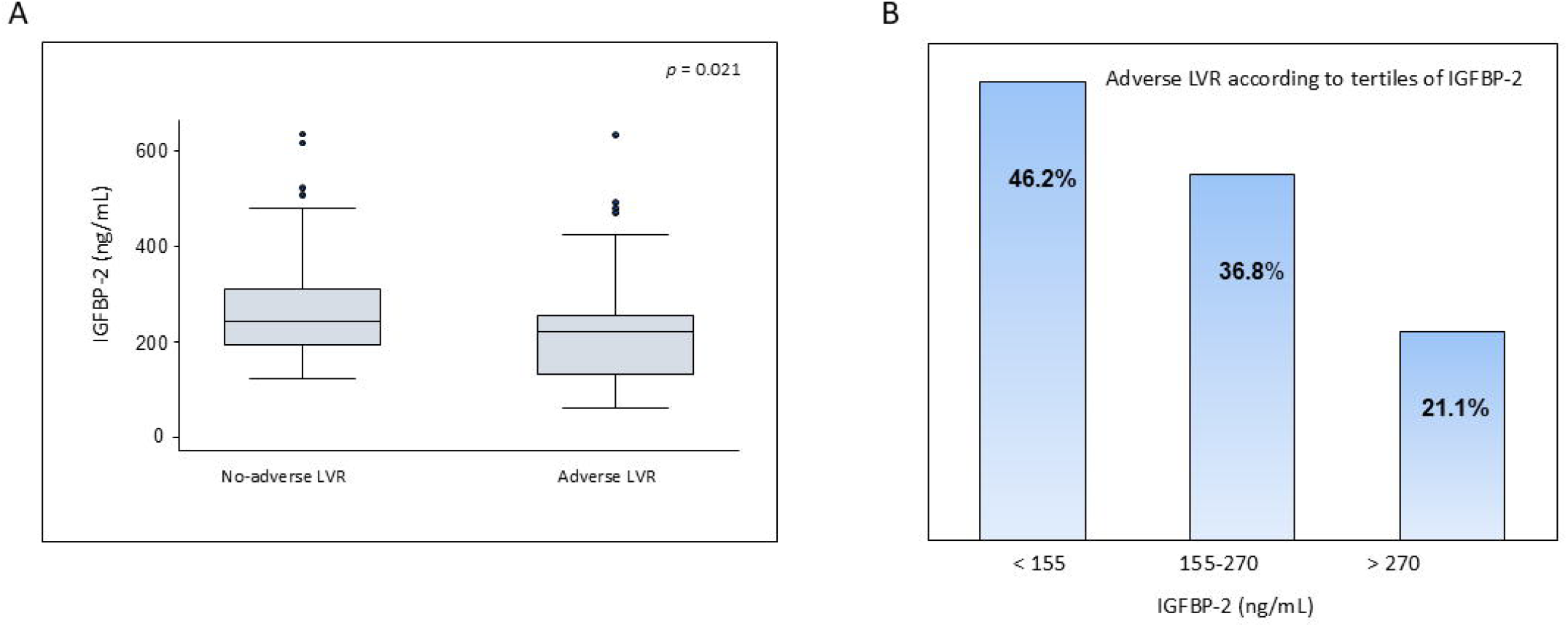
(A); Plasma peak baseline levels of IGFBP-2 in the study population measured. The box plots represent the median values (ng/mL) with interquartile range and the dots signify the outlier values plotted as individual points. *p* value is considered statistically significant when <0.05. (B); Percentage of adverse LVR patients according to tertiles of IGFBP-2 (ng/mL).

### ROC curve analysis and IGFBP-2–based risk prediction of adverse LVR following MI

Based on the variables that remained independently associated with adverse LVR in multivariate analyses (Table 2), we sequentially constructed nested models to assess their incremental predictive value. Model 1 included only ΔLVEF (change in LVEF from baseline to 12 months). Model 2 incorporated HDL-C in addition to ΔLVEF, and Model 3 combined ΔLVEF, HDL-C, and IGFBP-2. Likelihood-ratio (χ^2^) tests demonstrated that adding HDL-C to ΔLVEF significantly enhanced model performance (Model 2 vs. Model 1, χ^2^ = 9.10, p = 0.003), with a further significant gain achieved by adding IGFBP-2 (Model 3 vs. Model 2, χ^2^ = 4.72, *p*= 0.030). ROC curve analyses (Figure 2) revealed a progressive increase in predictive accuracy, with the area under the curve rising from 0.776 for Model 1 to 0.801 for Model 3, suggesting a meaningful trend toward improved discrimination (p _Model 3_ vs. _Model 1_ = 0.08).

**Figure 2.**
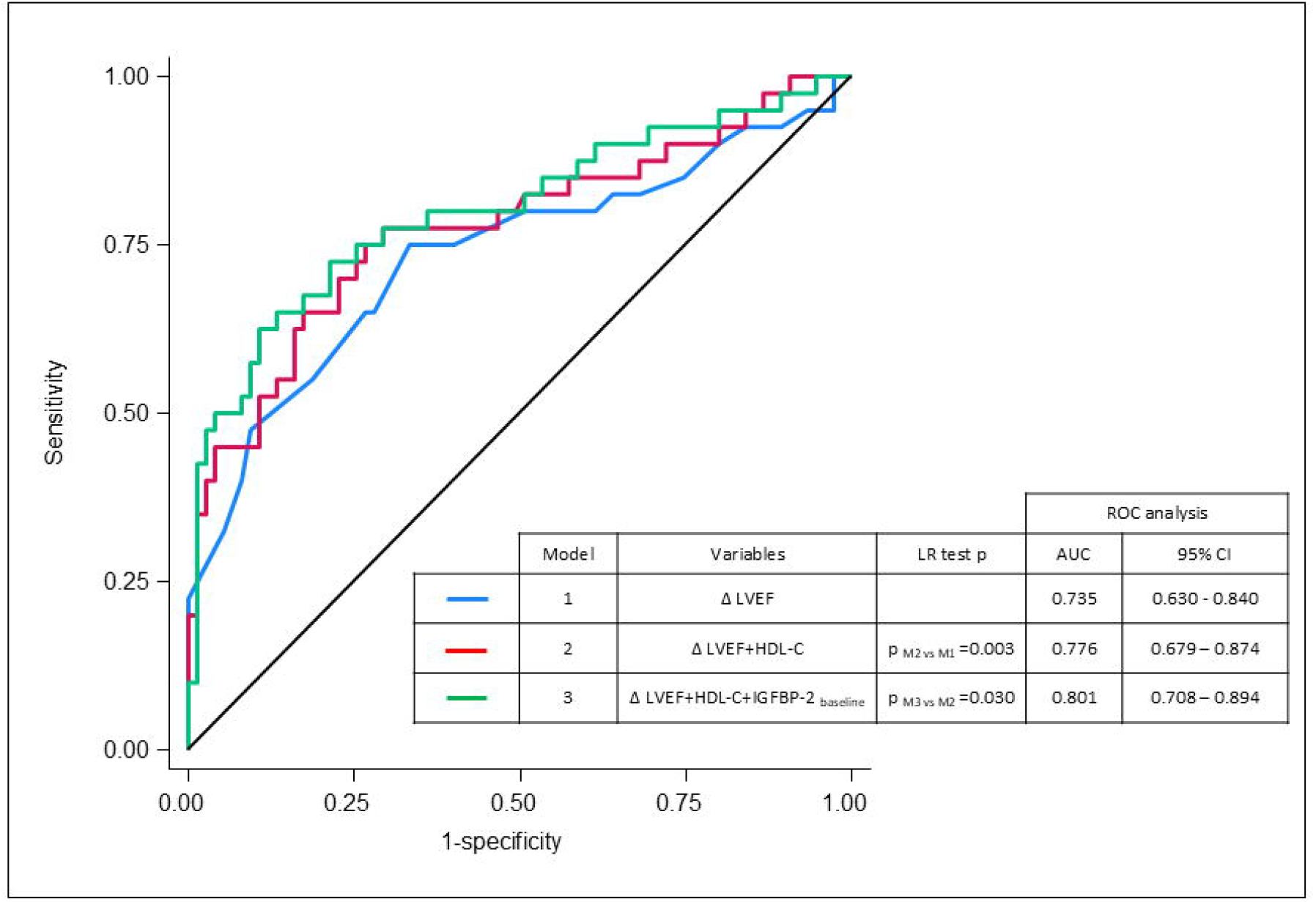
Receiver operating characteristic (ROC) curve analysis evaluating the performance of Model 1: ΔLVEF; Model 2: ΔLVEF+HDL-C and Model 3: ΔLVEF+HDL-C+ IGFBP-2. AUC, area under the curve; CI, confidence interval.

## Discussion

Although our cohort included a limited number of patients, it reflects real-world post-MI populations (14): 34% of patients developed adverse LVR at 12 months despite timely PCI and optimal medical therapy. This underscores the need for additional biomarkers to improve risk stratification after MI. Extending our previous work on biomarker trajectories underlying post-MI cardiac remodeling, we have now identified IGFBP-2 as a promising prognostic biomarker for adverse LVR after MI.

Lower baseline levels of IGFBP-2 were independently associated with an increased risk of adverse LVR, underscoring its role as a meaningful biomarker in the early risk stratification of post-AMI patients. IGFBP-2 is a secreted glycoprotein traditionally known for modulating the availability of insulin-like growth factors (IGFs), which are involved in cellular growth, metabolism, and tissue repair (15, 16). Beyond its IGF-dependent effects, IGFBP-2 also exerts IGF-independent actions, including anti-inflammatory, anti-fibrotic, and metabolic regulatory functions (15-17) but its role in post-MI cardiac remodeling remains poorly defined. IGFBP2 is highly expressed in a variety of tissues (e.g. liver, adipocyte, the reproduction and central nervous systems) and is dynamically regulated under stress and injury conditions. Interestingly, in our cohort, patients with adverse LVR had significantly lower IGFBP-2 levels, consistent with evidence that IGFBP-2 is inversely associated with insulin resistance and key components of the metabolic syndrome (18, 19). Thus, our findings suggest a link between insulin resistance, impaired myocardial repair, and a pro-inflammatory and pro-fibrotic cardiac environment—key drivers of maladaptive ventricular remodeling and HF progression. Furthermore, our data indicate that features of metabolic dysfunction—specifically hyperglycemia, hypertriglyceridemia, low HDL-C, and reduced circulating IGFBP-2—may contribute to the pathophysiological substrate underlying post-MI LVR. The clustering of these metabolic traits in the low IGFBP-2 group suggests that this protein may not only serve as a marker of cardiometabolic risk but could also represent a mechanistic link between metabolic dysfunction and maladaptive myocardial remodeling. IGF-independent actions of IGFBP-2 may contribute to adverse LVR through its Arg-Gly-Asp (RGD) motif that facilitates interaction with integrins, particularly αvβ3 and α5β1 (20). These interactions can activate intracellular signaling pathways, such as PI3K/Akt and FAK, which are involved in regulating cell adhesion, survival, and matrix remodeling. In the context of myocardial injury, downregulation of IGFBP-2 may impair these protective pathways, promoting apoptosis, extracellular matrix disorganization, and fibroblast activation—key hallmarks of maladaptive remodeling.

Although not reaching statistical significance, the trend observed in ROC curve analyses indicates that IGFBP-2 may provide incremental prognostic value and represents a promising candidate biomarker for inclusion in post-MI risk assessment models. It should be noted that IGFBP-2 has been identified as a prognostic biomarker for cardiovascular mortality in patients with HF (21). In addition, Muessig et al demonstrated in the CULPRIT-SHOCK trial its utility in risk stratification among patients with severe aortic stenosis undergoing transcatheter aortic valve implantation (22). Recently, post-hoc analysis of the CULPRIT-SHOCK trial demonstrated that IGFBP-2 levels are independently associated with higher 30-day and one-year mortality in patients with AMI complicated by cardiogenic shock, reinforcing its role as a metabolically linked biomarker with poor prognostic significance after AMI (23). Thus, identifying patients with low IGFBP-2 levels could help target metabolic interventions, such as insulin sensitizers, lipid-lowering agents, or anti-inflammatory therapies, in the early post-MI phase to potentially prevent or mitigate adverse LVR. The implications of these findings extend beyond risk stratification. The evolving landscape of HF management increasingly emphasizes metabolic modulation, as evidenced by the recent clinical success of SGLT2is and the emerging role of GLP-1 RAs. These agents exhibit pleiotropic effects that include favorable impacts on weight, insulin resistance, inflammation, and potentially myocardial remodeling. Recent evidence suggests that GLP-1 RAs may exert cardioprotective effects via both direct myocardial and systemic metabolic pathways (5, 24-26). Notably, the dual GIP and GLP-1 RA tirzepatide substantially increased IGFBP-2 levels in a dose and time-dependent pattern (27, 28). Recently, the SELECT trial (Semaglutide Effects on Cardiovascular Outcomes in People With Overweight or Obesity) demonstrated that semaglutide, a GLP-1 RA, significantly reduced major adverse cardiovascular events (MACE) in overweight or obese patients without diabetes (29) and composite HF endpoints in those with and without clinical HF, regardless of HF subtype (30). Since GLP-1 RA reduces systemic inflammation and improves cardiac outcomes, it’s plausible that improvements in IGFBP-2 levels could be associated with, or even mediate, some of the cardiac benefits observed in the SELECT trial. In this context, IGFBP-2 might serve as a biomarker not only for risk prediction but also for treatment response to such cardiometabolic therapies, paving the way for a precision medicine approach in post-MI care. Additionally, future investigations should explore whether therapeutic modulation of IGFBP-2 or its upstream regulators could mitigate LVR and HF development. In conclusion, IGFBP-2 emerges from this study as a novel, metabolically linked biomarker with independent prognostic value for adverse LVR following AMI. By bridging metabolic dysfunction and structural cardiac outcomes, IGFBP-2 could enrich current biomarker-based paradigms and inform future therapeutic interventions, particularly in an era where cardiometabolic comorbidity is central to HF pathogenesis and management.

### Study Limitations

Limitations should be acknowledged in the interpretation of our findings. First, although the study population was adequate to detect significant associations, the relatively small number of patients limits the statistical power for subgroup analyses and increases the risk of residual confounding. The observed trend to improvement in predictive performance with IGFBP-2 in ROC analyses, although not statistically significant, suggests a potential incremental role. Nevertheless, the borderline significance underscores the need for cautious interpretation, and validation in larger, independent cohorts will be essential to confirm the clinical utility of IGFBP-2 as an additive biomarker for risk stratification after MI. Second, the inclusion of both STEMI and NSTEMI patients introduced heterogeneity in clinical presentation. As NSTEMI patients typically present with smaller infarcts and less pronounced ventricular remodeling, this may have reduced biomarker-based associations and the ability to identify high-risk individuals within this subgroup.

## Conclusions and Clinical perspectives

The relationship of IGFBP-2 with metabolic dysregulation highlights its potential utility in guiding cardiometabolic risk stratification. For example, IGFBP-2–based models could support tailored surveillance strategies and serve as a potential marker of response to emerging therapies—such as GLP-1 RAs—that target metabolic pathways implicated in HF development. Future studies assessing longitudinal IGFBP-2 dynamics before and after GLP-1 RA initiation in post-MI patients, along with comparative studies versus placebo or standard care, may better define its role as a biomarker of therapeutic response and cardiac remodeling.

## Data availability

The data underlying this article will be shared on reasonable request to the corresponding author.

## Author’s contribution

All authors contributed to the article and approved the submitted version.

## Acknowledgements

We would like to express our sincere gratitude to the nurses, the patients and all investigators involved in this study, without whom the study would not have been possible. We thank Roche Diagnostics for generously providing the reagents for the GDF-15 assay. We are grateful to Dr AV. Cantero and D. Tayac for assistance in calibrating the GDF-15 assay, as well as the technical staff of the Clinical Biochemistry Laboratory at CHU Rangueil, Toulouse.

---

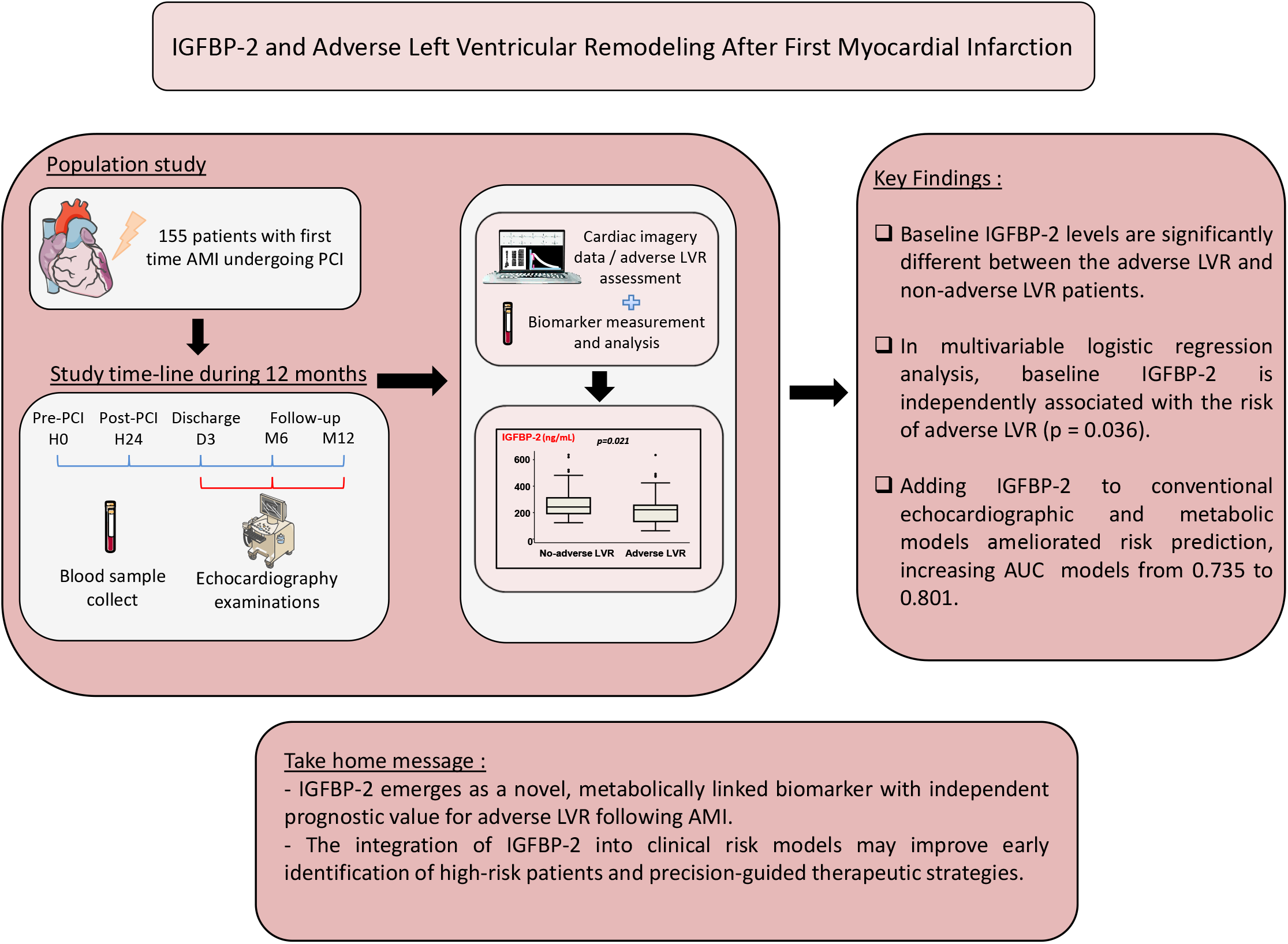

